# Impact of intermittent preventive treatment of malaria in pregnancy with sulfadoxine-pyrimethamine on sexually transmitted and reproductive tract infections: results from a randomised trial in Uganda

**DOI:** 10.1101/2025.07.02.25330769

**Authors:** Harriet Adrama, Erin J. Dela Cruz, Nida Ozarslan, Abel Kakuru, Bakar Odongo, Stephanie L. Gaw, Jade Benjamin-Chung, Jimmy Kizza, Miriam Aguti, John Ategeka, Peter Olwoch, Miriam Nakalembe, Bishop Opira, Tamara D. Clark, Moses R. Kamya, Philip J. Rosenthal, Grant Dorsey, Michelle E. Roh

## Abstract

**Background:** In sub-Saharan Africa, sexually transmitted and reproductive tract infections (STIs/RTIs) are important, but underdiagnosed risk factors for adverse pregnancy outcomes. Sulfadoxine-pyrimethamine (SP), used for intermittent preventive treatment of malaria in pregnancy (IPTp), may reduce STI/RTI burden due to its antibacterial activity. We assessed the impact of IPTp regimens on STI/RTI prevalence at delivery and associations between these infections and adverse birth outcomes.

**Methods:** We conducted a secondary analysis of a randomized controlled trial comparing monthly IPTp with SP, dihydroartemisinin-piperaquine (DP), or DP+SP among pregnant women in Uganda. Vaginal swabs collected at or near delivery were tested for *Chlamydia trachomatis*, *Neisseria gonorrhoeae*, *Trichomonas vaginalis*, and Group B *Streptococcus* (GBS) using GeneXpert; bacterial vaginosis was assessed using Nugent scoring. Log-binomial regression was used to compare STI/RTI prevalence by IPTp arm, using IPTp-DP as the reference arm. Multivariable Poisson regression with robust standard errors was used to evaluate associations between infections and preterm delivery, term low birthweight (LBW), overall LBW, and small-for-gestational age.

**Results:** Among the 2265 participants assessed, IPTp-SP reduced prevalence of *C. trachomatis* by 80% (2.5% vs. 12.4%; RR=0.20, 95% CI: 0.12-0.33) and of GBS by 35% (7.7% vs 11.7%; RR=0.65, 95% CI: 0.43-0.99) compared to IPTp-DP. *C. trachomatis* was associated with increased preterm delivery risk (RR=1.86, 95% CI: 1.07-3.25) and GBS was associated with increased term LBW risk (RR=2.08, 95% CI: 1.06-4.08).

**Conclusions:** Monthly IPTp-SP reduced the prevalence of *C. trachomatis* and GBS. These infections were associated with adverse birth outcomes, highlighting the potential non-malarial benefits of IPTp-SP.

**Key Messages:**

- In sub-Saharan Africa, management of sexually transmitted and reproductive tract infections (STIs/RTIs) relies on syndromic management, despite its high prevalence and potential risks associated with asymptomatic infections.
- Prior studies suggest that sulfadoxine-pyrimethamine (SP), the standard-of-care drug used for intermittent preventive treatment of malaria in pregnancy (IPTp), may exhibit activity certain STI/RTI pathogens, likely stemming from the sulfonamide component of the drug.
- Using data from a randomized trial comparing monthly IPTp regimens, we found IPTp-SP was associated with an 80% [95% CI: 67%-88%] reduction in *Chlamydia trachomatis* (2.5% versus 12.4%) and a 35% [95% CI: 1%-57%] reduction in Group B *Streptococcus* (7.7% vs. 11.7%) compared to IPTp-DP, an antimalarial with no known antibiotic activity.
- *C. trachomatis* was associated with an increased risk of preterm delivery (RR=1.86 [95% CI: 1.07-3.25]); Group B *Streptococcus* colonization was associated with an increased risk of term low birthweight (RR=2.08 [95% CI: 1.06-4.08]).
- IPTp-SP appears to offer benefits independent of malaria prevention through its effects on certain STI/RTIs pathogens, potentially contributing to a decrease in adverse birth outcomes. These findings are relevant as replacements to SP for IPTp are being considered.

## Introduction

In sub-Saharan Africa, the burdens of *Plasmodium falciparum* malaria, sexually transmitted infections (STIs), and reproductive tract infections (RTIs) are among the highest globally. Each year, an estimated 12.4 million pregnant women in the region are exposed to malaria infection,^1^ and 3 million are diagnosed with an STI or RTI.^2^ Most STIs and RTIs are asymptomatic, making them difficult to detect and treat, yet they can trigger inflammatory pathways that lead to adverse birth outcomes including stillbirth, preterm delivery, and low birthweight (LBW).^3^ To prevent malaria in pregnancy, the World Health Organization (WHO) recommends intermittent preventive treatment in pregnancy (IPTp), in which sulfadoxine-pyrimethamine (SP) is administered at scheduled intervals, from the second trimester of pregnancy until delivery.^4^ No comparable intervention exists for STI/RTI control. In sub-Saharan Africa, the WHO recommends routine screening only for syphilis and HIV infection, while other STIs/RTIs are managed syndromically,^5^ potentially missing an estimated 40–70% of infections.^6^

Over the past two decades, the spread of *P. falciparum* resistance to SP in Africa^7^ has motivated efforts to identify alternative IPTp regimens. The most extensively studied candidate is dihydroartemisinin-piperaquine (DP), a highly potent antimalarial with no known antibacterial activity. In a recent meta-analysis of six trials,^8^ IPTp-DP was markedly more effective than IPTp-SP at reducing malaria risk, but not at improving birth outcomes.^8^ This surprising result led to the hypothesis that SP may improve birth outcomes through mechanisms independent of its antimalarial activity. Several observational studies have supported this hypothesis; increasing doses of IPTp-SP were associated with decreasing LBW risk, even in areas with very low malaria transmission^9, 10^ or high-level *P. falciparum* resistance to SP.^11^

One possible mechanism underlying the non-malarial benefits of SP is its activity against STI/RTI pathogens. The sulfadoxine component of SP belongs to the sulfonamide class of antibiotics, which have demonstrated some *in vitro* activity against *Neisseria gonorrhoeae*, *Chlamydia trachomatis*, and Group B *Streptococcus* (GBS).^12–15^ Sulfonamides were previously used to treat *N. gonorrhoeae*^16, 17^ and *C. trachomatis*,^18^ but were replaced due to the rapid emergence of resistance. While various sulfonamides have been studied for their antimicrobial activity against STI/RTIs, few studies have directly evaluated SP.^12, 19^ In a recent randomized controlled trial, monthly IPTp-SP reduced *C. trachomatis* prevalence by 76% compared to IPTp-DP, and by 66% compared to IPTp-DP plus a single dose of azithromycin.^3^ However, the impact of IPTp-SP on GBS was not evaluated, despite prior *in vitro* evidence suggesting intermediate activity.^12^ Moreover, it is unclear whether SP’s potential activity on STIs/RTIs contribute to improved birth outcomes. We therefore evaluated the impact of IPTp-SP on STIs/RTI risk and the extent to which these effects may mediate adverse birth outcomes.

## Methods

### Study population and design

We conducted a secondary analysis of data from a three-arm, double-blinded, placebo-controlled randomized controlled trial (NCT04336189) evaluating monthly IPTp with SP, DP, or a combination of DP and SP (DP+SP) for the prevention of adverse birth outcomes.^20^ The trial was conducted in Busia District of southeastern Uganda, an area of intense, perennial malaria transmission.

Between December 2020 and December 2023, 2757 participants were enrolled. Eligibility criteria included: age ≥16 years, HIV-negative status, viable singleton pregnancy between 12-20 gestational weeks confirmed by ultrasound, and agreement to receive antenatal care and deliver at the study clinic. Participants were excluded if they reported a history of serious adverse events related to study drugs or previously received IPTp-SP in their current pregnancy. We further restricted our analyses to participants who had a vaginal swab collected.

Intervention randomization was conducted at a 1:1:1 ratio. Study drugs were initiated at 16 or 20 gestational weeks depending on the gestational age at enrolment and administered every four weeks until delivery. SP was administered as a single dose consisting of three tablets of 500 mg of sulfadoxine/25 mg pyrimethamine (Kamsidar, Kampala Pharmaceutical Industries). DP was administered as three consecutive daily doses; each dose consisted of three tablets of 40 mg dihydroartemisinin/320 mg piperaquine (Duo-Cotexcin, Holley-Cotec) given for three consecutive days. The first dose of study drugs was directly observed; the second and third were taken at home. Placebo pills were given to ensure all participants received the same number and colour of tablets for each IPTp course.

### Study procedures

At enrolment, participants underwent a standardized examination and were tested for syphilis. Women who tested positive for syphilis were treated with penicillin. Home visits were conducted to collect baseline data on household characteristics and malaria prevention measures. Routine antenatal care visits were scheduled every four weeks up to delivery for IPTp administration, a clinical examination, and to collect blood samples.

At delivery, standardized procedures were used to document delivery details, collect vaginal swabs, and assess newborn outcomes. Participants were strongly encouraged to deliver at the hospital adjacent to the study clinic; those delivering elsewhere were visited by study staff at or shortly after delivery. Vaginal swabs were rotated against the vaginal wall 2-3 times, rolled onto microscope slides to perform Nugent scoring for bacterial vaginosis testing, and placed into an XpertlZl Vaginal/Endocervical Specimen Collection Kit (SWAB/A-50, Cepheid, Sunnyvale, CA, USA) for storage at-80°C. If pre-delivery sample collection was not feasible (active labour or delivery outside of the study clinic), swabs were obtained at a postpartum visit, ranging from 6-45 days after delivery. Additional details of study procedures are described elsewhere.^20^

### STI/RTI diagnosis

Vaginal swabs were tested for *C. trachomatis*, *N. gonorrhoeae*, *T. vaginalis*, and GBS using GeneXpert qPCR (XpertlZl CT/NG, XpertlZl TV, and XpertlZl Xpress GBS; Cepheid). Due to budgetary constraints, GBS testing was discontinued midway through the trial. Bacterial vaginosis assessment was performed using Nugent scoring by trained, blinded microscopists; a score of 7-10 indicated a positive diagnosis.^21, 22^

### Birth outcomes

Birth outcomes of interest included: preterm delivery (delivery <37 gestational weeks), LBW (birthweight <2500 grams), term LBW (LBW among infants born ≥37 gestational weeks), and small-for-gestational age (SGA; <10th percentile of birthweight-for-gestational age based on INTERGROWTH-21st standards^23^). Preterm delivery included live births and foetal losses. Due to missing birthweights for foetal losses, LBW and SGA-based outcomes were restricted to live births. Stillbirths and miscarriages were not evaluated as separate outcomes due to the rarity of these outcomes (<2%).

### Covariates

We used a directed acyclic graph (DAG) to guide our decisions on covariate adjustment (**Figure 1**). Maternal age, mid-upper arm circumference, weight, and gravidity at enrolment were modelled as continuous variables. Maternal education was categorized into three levels, and socioeconomic status was measured as household wealth tertiles, derived using principal components analysis of common household items.^24^ Current or recent alcohol use and history of any adverse delivery outcome (preterm delivery, miscarriage, or stillbirth) were measured as binary variables at enrolment.

**Figure 1.**
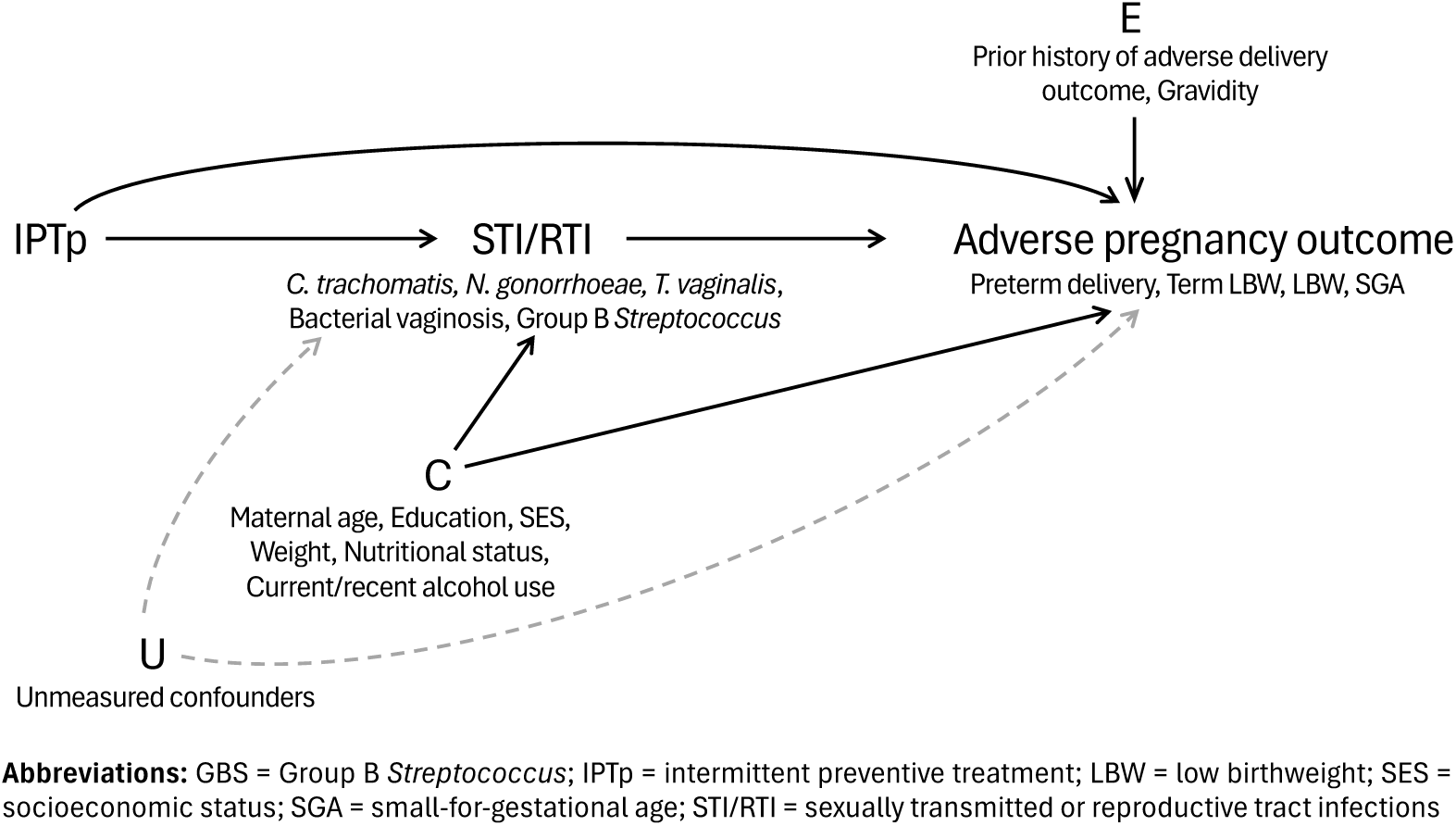
Directed acyclic graph (DAG) of the presumed relationship between IPTp regimens, STI/RTI risk, and adverse birth outcomes. C indicates the vector of measured covariates that we assumed were confounders of the STI-adverse birth outcome relationship. E includes the vector of covariates that may not meet the definitions of a confounder (i.e., common cause of the exposure and outcome), but were important prognostic variables of the outcome. We expect that inclusion of these baseline covariates would not bias our effect estimates but may improve statistical precision. Grey dashed lines indicate potential causal pathways of how unmeasured variables (U) may confound the STI-adverse pregnancy outcome relationship.

### Statistical analysis

This study had three objectives: (1) evaluate whether monthly IPTp-SP reduced the prevalence of STIs/RTIs compared to IPTp-DP, (2) determine associations between STIs/RTIs and adverse birth outcomes, and (3) for STIs/RTIs associated with both SP exposure and adverse birth outcomes, estimate the extent to which the overall effect of IPTp-SP on birth outcomes was mediated through its effects on STIs/RTIs. For the first objective, we conducted intention-to-treat analyses based on randomized IPTp assignment. We used IPTp-DP as the reference arm, assuming that DP has no effect on STIs/RTIs, and compared it with IPTp-SP, IPTp-SP+DP, and the combined IPTp-SP and IPTp-SP+DP arms. Log binomial regression was used to estimate unadjusted relative risk ratios (RRs) and 95% confidence intervals (CIs). Risk differences (RDs) and the number needed to treat (NNT) were computed using the `margins` command in Stata.

To determine associations between STIs/RTIs and adverse birth outcomes, we fit multivariable Poisson regression models with robust standard errors. Each model included the STI/RTI of interest as the primary exposure and adjusted for IPTp arm, household wealth, maternal age, weight, mid-upper arm circumference, education, current/recent alcohol use, history of adverse delivery outcome, and gravidity. Continuous variables were modelled as three-knot restricted cubic splines.

Lastly, we used causal mediation analyses to estimate the effect of IPTp-SP on birth outcomes mediated through STI/RTI prevention. Analyses were conducted using the `CMAverse` R package^25^ using parametric g-formula.^26^ To ensure mediation analyses focused on biologically and statistically plausible pathways, we limited our analyses to STI/RTIs that demonstrated associations with both the exposure (IPTp-SP) and the outcome (birth outcomes). These decisions were guided by the magnitude of estimated effects, supported by 95% CIs that excluded null effects. Logistic regression was used for mediator and outcome models and included an exposure-mediator interaction term, regardless of a statistically significant interaction.^27^ We report the total (overall) effect and the natural indirect (mediated) and direct (non-mediated) effects, which sets the mediator value to what it would have naturally taken had participants been assigned to IPTp-DP and IPTp-SP, respectively. Mediation analyses excluded the IPTp-DP+SP arm to ensure comparisons were not affected by drug-drug interactions.^28^ Percentile bootstrapped 95% CIs were computed based on 1000 simulations.

All analyses were performed using Stata 16.1 (Stata Corp, College Station, Texas) and R (version 4.4.0, R Project for Statistical Computing, Vienna, Austria).

### Sensitivity analyses

Sensitivity analyses were conducted to assess the robustness of estimated effects. First, although vaginal swabs for STI/RTI testing were intended to be collected at delivery, 28% (n=705) were obtained at delivery, 59% (n=1509) obtained postpartum, and 13% (n=324) were either missing or collected with unknown timing. Participant characteristics differed between those sampled at delivery versus postpartum, including IPTp assignment, STI/RTI status, and birth outcomes (Appendix 1). To avoid differential misclassification or selection bias, we treated non-delivery samples as missing and performed multiple imputation (MI) using chained equations using the `MISL` R package,^29^ which employs an ensemble machine learning approach (SuperLearner) to flexibly model relationships. Model predictors included IPTp arm, maternal mid-upper arm circumference, age, weight, household wealth index, and preterm delivery. Analyses were computed on each imputed dataset and pooled using Rubin’s combining rules.^30^ We also used inverse probability of selection weighting (IPSW) to reweight participants with delivery samples to approximate the full trial population. Additional methodological details are provided in Appendix 2.

Second, causal interpretation of our estimated effects of STIs/RTIs on birth outcomes and mediation analyses relies on the assumption of no unmeasured confounding. While this assumption likely holds for IPTp-SP effects due to treatment randomization, it may not for STI/RTI-outcome associations. We therefore report E-values^25, 31, 32^ alongside estimates to quantify the minimum strength that an unmeasured confounder would need to have with both the exposure and outcome, conditional on measured covariates, to fully explain observed effects.

## Results

Among the 2757 participants enrolled in the trial, 2538 (92%) were followed through delivery, and 2265 (82%) had a vaginal swab collected (**Figure 2**). Of these 2265 swabs, 705 (31%) were collected prior to delivery, 1509 (66.7%) were collected postpartum, and for 51 (2.3%) the time of collection was not recorded. Among samples collected postpartum, the mean time from delivery to collection was 28 days (standard deviation [SD]: 5). The mean age at enrolment was 24.7 years (SD: 6.3), 96% completed primary school, and 8.5% reported a prior adverse birth outcome. Fourteen participants (0.6%) tested positive for syphilis at enrolment and were treated. Baseline characteristics were balanced across IPTp arms (**Table 1**).

**Figure 2.**
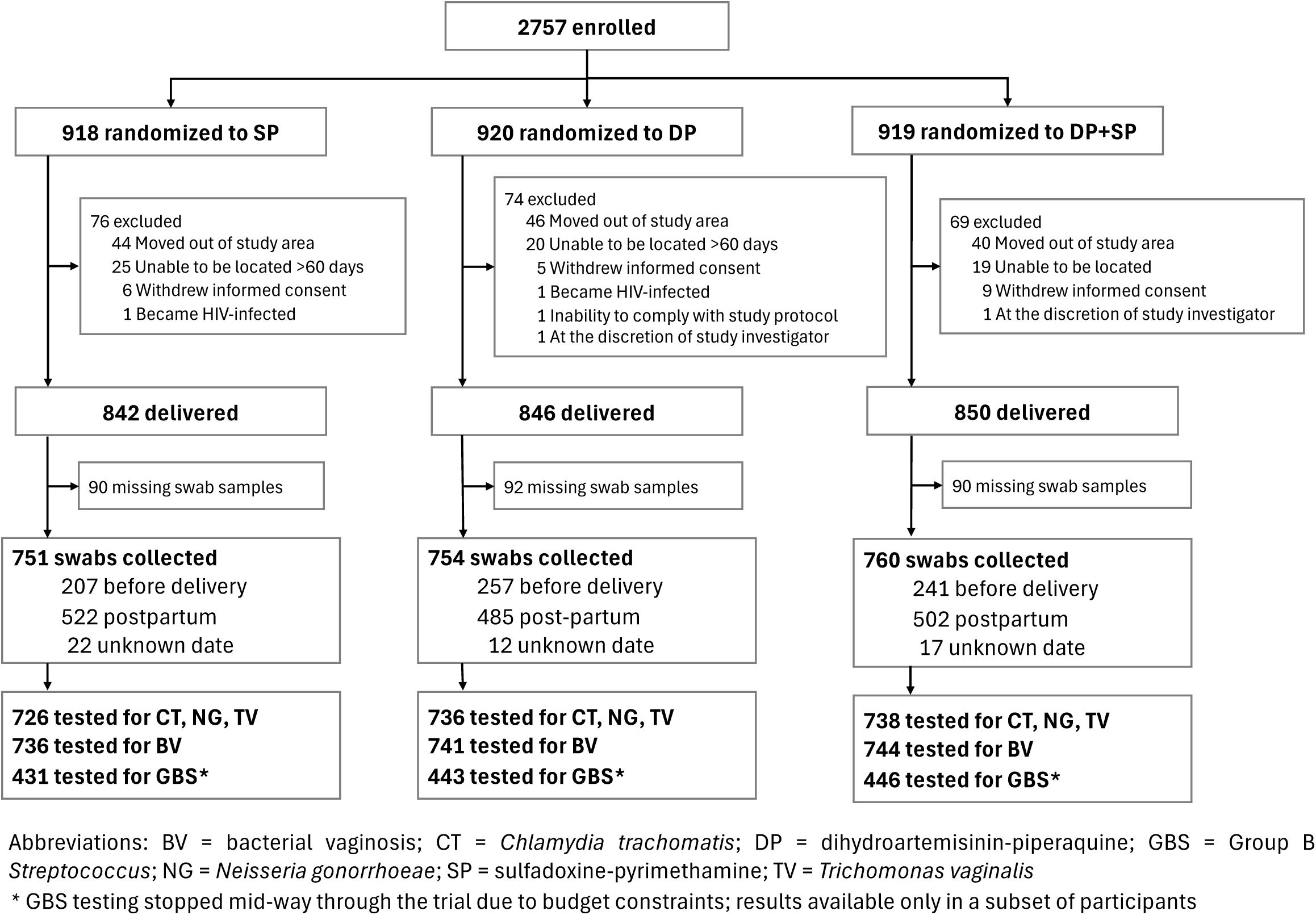
Study participant flowchart.

**Table 1.** Baseline characteristics of trial participants included in the final analytic sample.

| **Characteristics** | **SP**<br>**N = 751** | **DP**<br>**N = 754** | **DP+SP**<br>**N = 760** |
| --- | --- | --- | --- |
| Maternal age in years, n (%) |  |  |  |
| 16-20 | 230 (30.6%) | 227 (30.1%) | 257 (33.8%) |
| 21-25 | 205 (27.3%) | 223 (29.6%) | 211 (27.8%) |
| 26-30 | 178 (23.7%) | 142 (18.8%) | 157 (20.7%) |
| 31-35 | 87 (11.6%) | 109 (14.5%) | 94 (12.4%) |
| 36+ | 51 (6.8%) | 53 (7.0%) | 41 (5.4%) |
| Gestational age at enrolment, n (%) |  |  |  |
| 12 to <16 | 415 (55.3%) | 424 (56.2%) | 379 (49.9%) |
| 16 to 20 | 336 (44.7%) | 330 (43.8%) | 381 (50.1%) |
| Gravidity, n (%) |  |  |  |
| Primigravidae | 177 (23.6%) | 188 (24.9%) | 218 (28.7%) |
| Secundigravidae | 167 (22.2%) | 157 (20.8%) | 151 (19.9%) |
| Multigravidae | 407 (54.2%) | 409 (54.2%) | 391 (51.4%) |
| Positive for syphilis, n (%) | 4 (0.5%) | 7 (0.9%) | 2 (0.3%) |
| Maternal weight (kg), mean (SD) | 57.1 (9.4) | 57.7 (9.9) | 57.2 (9.0) |
| Maternal height (cm), mean (SD) | 159.0 (6.2) | 159.1 (6.8) | 159.3 (6.0) |
| Maternal MUAC (cm), mean (SD) | 25.8 (2.8) | 25.9 (2.8) | 25.8 (2.7) |
| Maternal BMI, mean (SD) |  |  |  |
| <18.5 | 49 (6.5%) | 47 (6.2%) | 30 (3.9%) |
| 18.5 to <25 | 557 (74.2%) | 560 (74.4%) | 599 (78.8%) |
| 25 to <30 | 122 (16.2%) | 110 (14.6%) | 107 (14.1%) |
| 30+ | 23 (3.1%) | 36 (4.8%) | 24 (3.2%) |
| Household wealth index tertiles, n (%) |  |  |  |
| Lowest | 254 (34.0%) | 239 (31.9%) | 253 (33.4%) |
| Middle | 263 (35.2%) | 264 (35.2%) | 247 (32.6%) |
| Highest | 231 (30.9%) | 247 (32.9%) | 257 (33.9%) |
| Highest level of education, n (%) |  |  |  |
| None | 40 (5.3%) | 26 (3.4%) | 32 (4.2%) |
| Primary | 486 (64.7%) | 479 (63.5%) | 505 (66.4%) |
| ≥ O level | 225 (30.0%) | 249 (33.0%) | 223 (29.3%) |
| Current/recent alcohol use, n (%) | 35 (4.7%) | 35 (4.6%) | 28 (3.7%) |
| History of an adverse delivery outcome, n (%) | 74 (9.9%) | 59 (7.8%) | 57 (7.5%) |

### Effect of IPTp regimen on STI/RTI risk

In the IPTp-DP arm (reference), the prevalence of *C. trachomatis*, *N. gonorrhoeae*, *T. vaginalis*, bacterial vaginosis, and GBS colonization at or near delivery was 12.4%, 2.7%, 8.3%, 43.6%, and 11.7%, respectively. Compared to IPTp-DP, IPTp-SP had markedly lower prevalence of *C. trachomatis* (2.5% vs. 12.4%), corresponding to a RR=0.20 [95% CI: 0.12-0.33]), RD=-9.9% [95% CI:-12.5%,-7.3%], and NNT=10 [95% CI: 8, 14] (**Figure 3**). Similar reductions were observed when IPTp-DP was compared to the IPTp-DP+SP arm (RR=0.26 [95% CI: 0.17-0.41]) or to the combined IPTp-SP and IPTp-DP+SP arms (RR=0.23 [95% CI: 0.16-0.33]).

**Figure 3.**
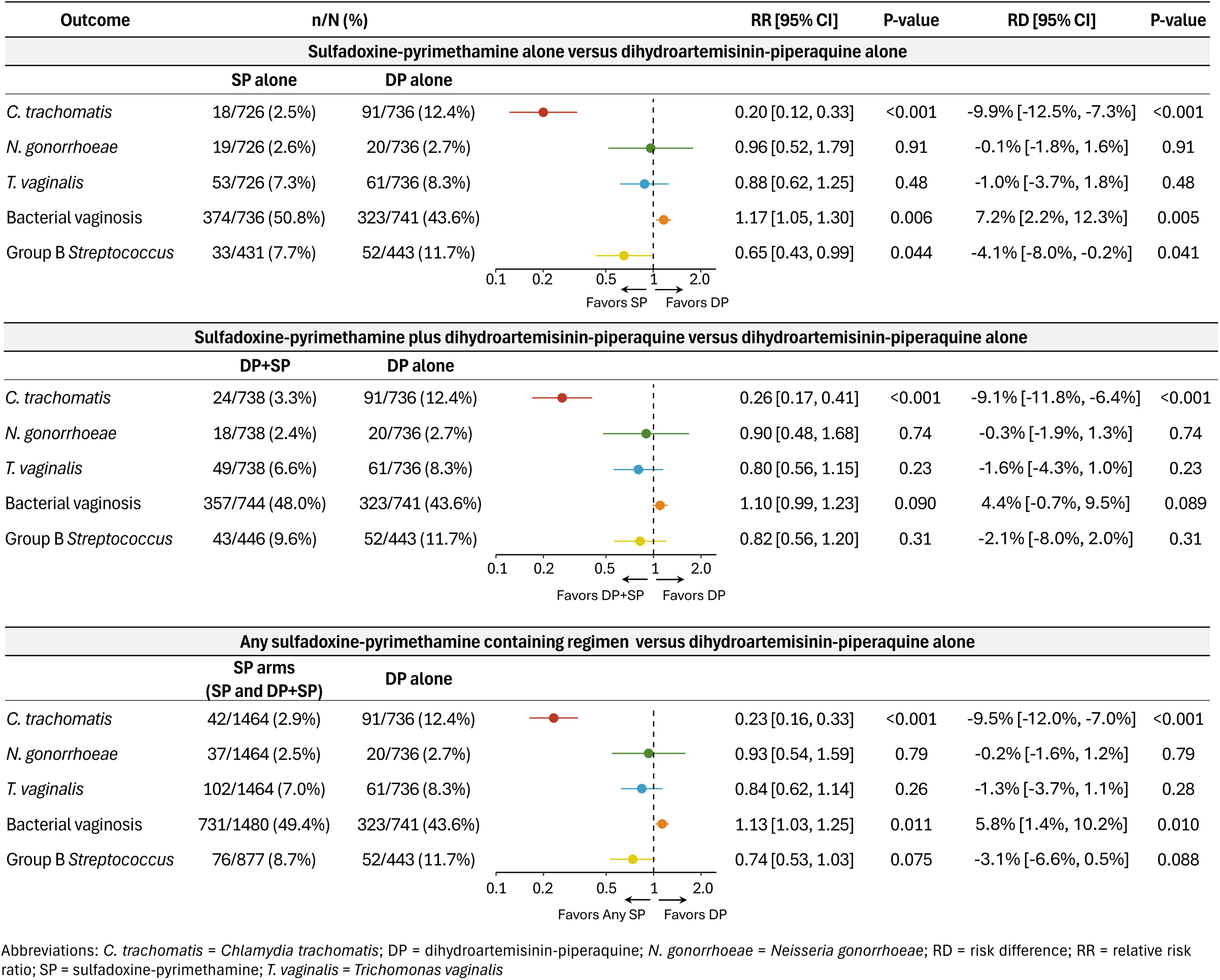
Effect of IPTp regimens on STI/RTI prevalence. Analyses were conducted using unadjusted log binomial regression models.

GBS prevalence was lower among participants randomized to SP-containing IPTp regimens compared to IPTp-DP (7.7% vs. 11.7%, any SP: RR=0.74 [95% CI: 0.53-1.03]; NNT=32 [95% CI: 15, 200]), with the largest reduction in the IPTp-SP arm (RR=0.65 [95% CI: 0.43-0.99]; NNT=24 [95% CI: 13, 500]). Notably, the prevalence of bacterial vaginosis was higher in the IPTp-SP arm (RR=1.17 [95% CI: 1.05-1.30]) and the IPTp-DP+SP arm (RR=1.10 [95% CI: 0.99-1.23]) relative to IPTp-DP (**Figure 3**). Effect sizes for *N. gonorrhoeae* and *T. vaginalis* were modest, with wide CIs that included the null.

Sensitivity analyses accounting for timing of swab collection were generally consistent with the primary results, with the exception of IPTp effects on bacterial vaginosis. For this analysis, MI and IPSW estimates were closer to the null (RR_MI_=1.09 [95% CI: 0.88-1.35] and RR_IPSW_=1.09 [95% CI: 0.85-1.40]) compared to the primary estimate (RR=1.17 [95% CI: 1.05-1.30]), (Appendix 3).

### Effect of STIs/RTIs on birth outcomes

In adjusted models, both *C. trachomatis* (RR=1.86 [95% CI: 1.07-3.25]) and bacterial vaginosis (RR=1.48 [95% CI: 1.07-2.05]) were associated with an increased risk of preterm delivery (**Figure 4**). Although point estimates suggested that *N. gonorrhoeae*, *T. vaginalis*, and GBS were also associated with an increased risk of preterm delivery, wide confidence intervals spanned both protective and harmful effects. For LBW-based outcomes, associations were generally null and highly imprecise, except for GBS, which was associated with an approximately two-fold increase in term LBW (RR=2.08 [95% CI: 1.06-4.08]) and LBW overall (RR=1.66 [95% CI: 0.94-2.94]). There was no clear evidence of an association between any STI/RTI and SGA risk.

**Figure 4.**
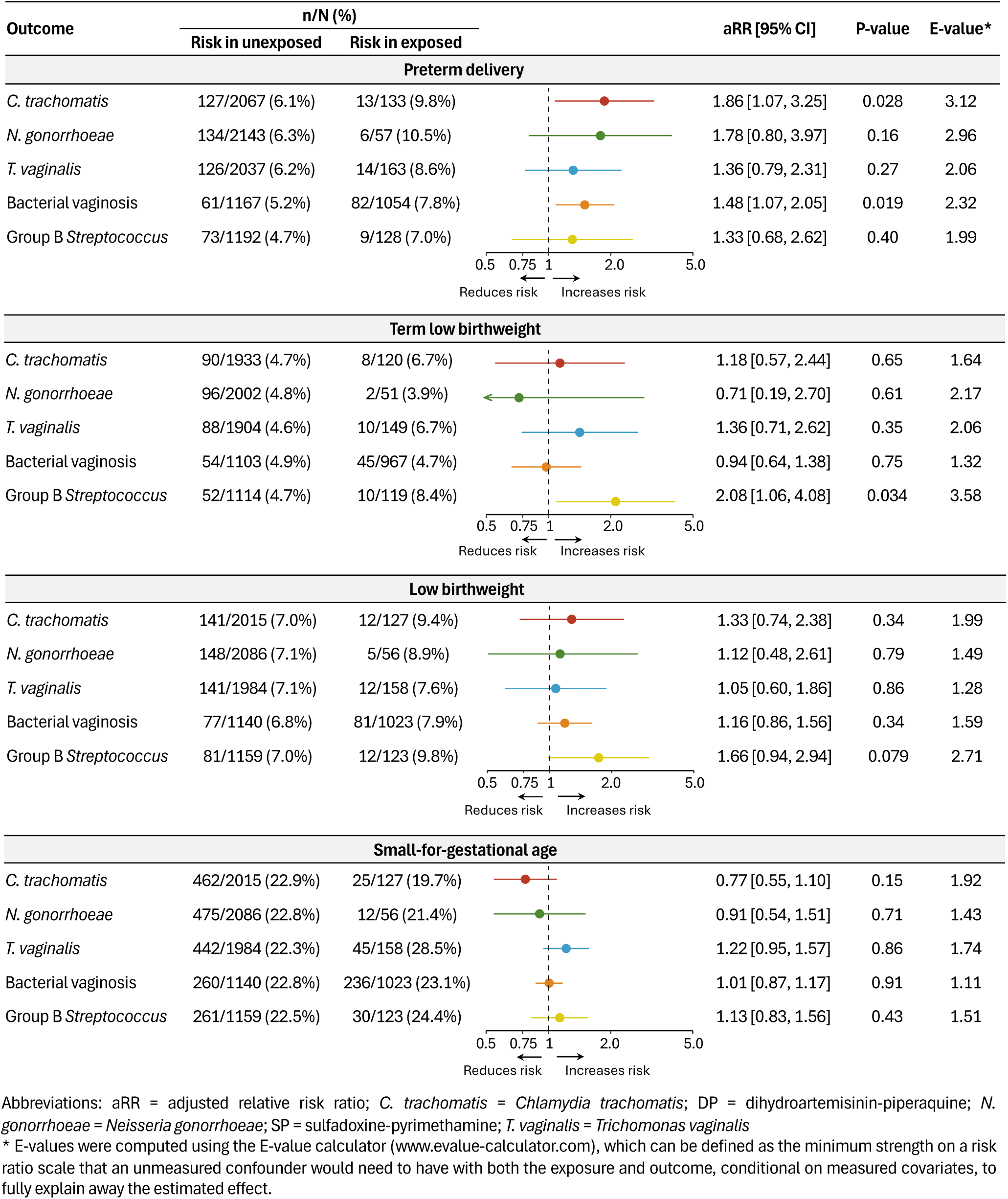
Associations between STIs/RTIs and adverse birth outcomes. Analyses were conducted using multivariable Poisson regression with robust standard errors, adjusting for IPTp arm, household wealth, maternal age, weight, mid-upper arm circumference, education, current/recent alcohol use, history of adverse delivery outcome, and gravidity.

E-values indicated that, for estimates with 95% CIs that excluded the null, an unmeasured confounder would need to have a RR≥2.0 with both the exposure and outcome, conditional on measured covariates, to fully account for the observed effects (**Figure 3**). Sensitivity analyses accounting for timing of swab collection were consistent with primary analyses (Appendix 3); MI estimates were generally attenuated toward the null, although 95% CIs for both MI and IPSW models overlapped with the primary results.

### Mediation analyses

Mediation analyses evaluated whether the effects of IPTp-SP on *C. trachomatis* and GBS contributed to its overall effects on preterm delivery and term LBW, respectively (**Table 2**). Compared to IPTp-DP, IPTp-SP was associated with an increased risk of preterm delivery (total effect: RR=1.39 [95% CI: 0.86-2.13]), however, this effect was partially offset by IPTp-SP’s greater prevention of *C. trachomatis* compared to IPTp-DP (indirect effect: RR=0.78 [95% CI: 0.57-1.03]). In contrast, IPTp-SP was associated with lower risk of term LBW compared to IPTp-DP (total effect: RR=0.28 [95% CI: 0.11-0.56]), but this effect was minimally mediated through its greater effect on preventing GBS (indirect effect: RR=0.97 [95% CI: 0.89-1.03]; % mediated: 1% [95% CI:-1%, 5%]). E-values for the indirect effects (1.82 for *C. trachomatis* and 1.14 for GBS), suggested moderate robustness to unmeasured confounding for the *C. trachomatis* estimate, but not GBS.

**Table 2.** Total, indirect, and direct effects* of IPTp-SP versus IPTp-DP on birth outcomes mediated by STI/RTI pathogens.

| STI/RTI | Outcome | Total Effect | Total NIE | Pure NDE | % Mediated<sup>^</sup> | E-value<sup>†</sup> |
| --- | --- | --- | --- | --- | --- | --- |
| *C. trachomatis* | Preterm delivery | 1.39 [0.86, 2.13] | 0.78 [0.57, 1.03] | 1.78 [1.03, 2.98] | -- | 1.82 |
| Group B *Streptococcus* | Term LBW | 0.28 [0.11, 0.56] | 0.97 [0.89, 1.03] | 0.28 [0.12, 0.55] | 1% [-1%, 5%] | 1.17 |

Abbreviations: *C. trachomatis* = *Chlamydia trachomatis*; LBW = low birthweight; NDE = natural direct effect; NIE = natural indirect effect; STI/RTI = sexually transmitted or reproductive tract infection
\* Analyses restricted to participants in the IPTp-SP and IPTp-DP arms only; the IPTp-DP+SP arm was excluded from mediation models to ensure comparisons were not affected by drug-drug interactions.
<sup>^</sup> Defined as the % of the total effect of the exposure on the outcome that is mediated through the specified mediator; calculated as the indirect effect divided by the total effect. % mediated not calculated if direct and indirect effects had opposing signs.
<sup>†</sup> Defined as the minimum strength on a risk ratio scale that an unmeasured confounder would need to have with both the mediator and outcome, conditional on measured covariates, to fully explain away the indirect effect.

## Discussion

In this randomised controlled trial comparing monthly IPTp regimens, IPTp-SP reduced the prevalence of *C. trachomatis* by 80% and GBS by 35% compared to IPTp-DP, with similar reductions observed for IPTp-DP+SP. These effects are noteworthy, as *C. trachomatis* was associated with a 1.86-fold increase in preterm delivery and GBS with a 2.08-fold increase in term LBW. Consistent with prior *in vitro* and clinical studies,^3, 12^ IPTp-SP did not reduce the prevalence of *N. gonorrhoeae* or *T. vaginalis* infection. Notably, IPTp-SP was associated with an increased risk of bacterial vaginosis compared to IPTp-DP, though this effect was attenuated in sensitivity analyses that considered the timing of sample collection. This result is consistent with prior studies which have found the vaginal microbiota shifts to a more diverse, bacterial vaginosis-like community shortly after delivery and in the early postpartum period.^33, 34^ Together, these findings suggest that, in addition to its role as an antimalarial, IPTp-SP reduces the risk of certain STI/RTI pathogens, with downstream impacts on adverse birth outcomes.

Mediation analyses further revealed that, although IPTp-DP had a stronger overall effect than IPTp-SP on reducing preterm birth, likely due to its more potent antimalarial activity,^8, 20^ IPTp-SP’s greater effect against *C. trachomatis* partially offset this effect. While IPTp-SP also reduced GBS colonization, this reduction was modest in absolute terms, and consequently, GBS mediated only a small fraction of IPTp-SP’s greater effect on term LBW compared to IPTp-DP, suggesting other mechanisms likely contribute to the observed benefit. These analyses suggest that IPTp regimens affect pregnancy outcomes through multiple biological pathways. Therefore, future research on alternative IPTp regimens should prioritize drug combinations that provide both effective malaria protection and broader maternal and neonatal health benefits.

In most of sub-Saharan African, STI/RTI screening during pregnancy relies on syndromic management due to limited diagnostic capacity,^5^ leading to undertreatment and likely excess adverse pregnancy outcomes. A modelling study from South Africa estimated that 25.5% of HIV-negative and 34.6% of HIV-positive pregnant women had undiagnosed STIs, for which treatment could have prevented up to 6.3% of preterm deliveries, 10.1% of stillbirths, and 9.1% of LBW infants.^35^ Our findings suggest that IPTp-SP may be a low-cost chemoprevention strategy to protect against *C. trachomatis* and GBS, which were present in 11-12% of women in our study and 14-16% in parts of sub-Saharan Africa.^36, 37^ Notably, the NNT to prevent one *C. trachomatis* infection was 10—a compelling figure given its associations with adverse pregnancy outcomes and neonatal complications, including conjunctivitis and pneumonia.^38^ The 35% reduction in GBS was similarly noteworthy, as this pathogen is not routinely screened for in resource-limited settings despite its links to prevent neonatal sepsis, which can have significant impacts on neonatal morbidity and mortality.^39^ Thus, even amid rising *P. falciparum* antifolate resistance, IPTp-SP effects on STIs/RTIs should be considered for its maternal and neonatal benefits beyond malaria prevention.

Our analysis had several limitations. First, while most swabs were collected postpartum (∼28 days post-delivery) rather than at delivery as planned, our sensitivity analyses accounting for STI/RTI measurement timing produced similar findings (except for bacterial vaginosis results). Second, estimates of STI/RTI-birth outcome associations and mediation analyses may be subject to unmeasured confounding, particularly related to sexual behaviour or concurrent antibiotic use. However, sensitivity analyses suggested that an unmeasured confounder would need to have a moderately strong association (RR≥2.0) with both the mediator and outcome to fully explain observed effects. Third, STI/RTI assessment was limited to select pathogens and did not include the broader spectrum of reproductive tract microbes or pathobionts potentially affected by IPTp-SP. Fourth, we did not assess chorioamnionitis or neonatal sepsis, which have been strongly linked to STI/RTIs but were not reliably measured in our study. Finally, we assumed IPTp-DP had no antibacterial activity. If it did, our estimates of IPTp-SP’s effect on STIs/RTIs may have been underestimated.

The WHO currently recommends IPTp in 34 malaria-endemic African countries, with SP remaining the standard-of-care despite rising antifolate resistance.^1, 40^ Our findings suggest SP reduces *C. trachomatis* by 80% and GBS by 35%, which could partially explain its positive impacts on birth and neonatal outcomes independent of its effects on malaria. As alternative IPTp regimens are being considered, these ancillary effects should be appreciated, especially in settings where access to STI/RTI screening and treatment remains limited.

## Declarations

## Ethics approval

The study was approved by the Makerere University School of Biomedical Sciences Research Ethics Committee (SBS 714), the Uganda National Council for Science and Technology (HS 2746), the Uganda National Drug Authority (CTC 0135/2020), and the University of California San Francisco Human Research Protection Program (19-29105).

## Data availability

Data can be made available upon reasonable request to the corresponding author and will be granted with approval by Principal Investigators, Grant Dorsey, Philip Rosenthal, and Moses R. Kamya.

## Supplementary data

Supplementary data are available at *IJE* online.

## Author contributions

AK, PJR, MRK, MER, and GD conceived the idea for the trial. HA, AK, JK, MA, JA, MN, BO, and TDC were responsible for implementing the trial and managing study participants. HA, PO, OB, and JA performed laboratory analyses and contributed to data acquisition. MER performed the statistical analysis with inputs from HA, EJD, NO, GD, and JBC. HA, EJD, NO, and MER wrote the first draft of the manuscript, with significant inputs from SLG, PJR, and GD. All authors interpreted the data and critically reviewed the manuscript.

## Use of artificial intelligence (AI) tools

AI tools were used solely for improving the readability of the manuscript and English grammar.

## Acknowledgements

We thank the trial participants, the dedicated staff at Masafu General Hospital, and the staff of the Infectious Diseases Research Collaboration and the University of California, San Francisco for their administrative and regulatory support.

