## Appendix for "Impact of intermittent preventive treatment of malaria in pregnancy with sulfadoxine-pyrimethamine on sexually transmitted and reproductive tract infections: results from a randomised trial in Uganda"

**Supplementary Appendix 1.** Characteristics of participants with STI/RTI results collected at delivery versus postpartum.

| **Characteristics** | **Sampled before delivery**  **(n=705)** | **Sampled postpartum or missing (n=1833)** | **p-value** |
| --- | --- | --- | --- |
| ***Enrollment characteristics*** | | | |
| Treatment arm, n/N (%) |  |  | 0.026 |
| SP | 207/705 (29%) | 635/1833 (35%) |  |
| DP | 257/705 (36%) | 589/1833 (32%) |  |
| DP+SP | 241/705 (34%) | 609/1833 (33%) |  |
| Gravidity, n/N (%) |  |  | 0.18 |
| Primigravida | 194/705 (28%) | 456/1833 (25%) |  |
| Secundigravida | 136/705 (19%) | 408/1833 (22%) |  |
| Multigravida | 375/705 (53%) | 969/1833 (53%) |  |
| Gestational age at enrolment in weeks, n/N (%) |  |  | 0.15 |
| ≤16 weeks | 411/705 (58%) | 1011/1833 (55%) |  |
| >16 weeks | 294/705 (42%) | 822/1833 (45%) |  |
| Maternal age in years, n/N (%) |  |  | 0.67 |
| <18 | 92/705 (13%) | 228/1833 (12%) |  |
| ≥18 to <25 | 304/705 (43%) | 766/1833 (42%) |  |
| ≥25 | 309/705 (44%) | 839/1833 (46%) |  |
| Household wealth tertiles, n/N (%) |  |  | 0.35 |
| Poorest | 248/703 (35%) | 591/1821 (32%) |  |
| Middle | 237/703 (34%) | 623/1821 (34%) |  |
| Least poor | 218/703 (31%) | 607/1821 (33%) |  |
| Maternal BMI in kg/m^2^, n/N (%) |  |  | 0.93 |
| <18.5 | 39/704 (6%) | 100/1833 (5%) |  |
| ≥18.5 to <25 | 531/704 (75%) | 1399/1833 (76%) |  |
| ≥25 to <30 | 106/704 (15%) | 270/1833 (15%) |  |
| ≥30 | 28/704 (4%) | 64/1833 (3%) |  |
| Highest level of education, n/N (%) |  |  | 0.35 |
| None | 26/705 (4%) | 78/1833 (4%) |  |
| Primary school | 448/705 (64%) | 1206/1833 (66%) |  |
| Secondary school and above | 231/705 (33%) | 549/1833 (30%) |  |
| Maternal MUAC in cm, mean (SD) | 25.7 (2.8) | 25.9 (2.7) | 0.039 |
| Current or recent alcohol use, n/N (%) | 82/1,830 (4%) | 33/705 (5%) | 0.83 |
| History of an adverse birth outcome,^#^ n/N (%) | 153/1,833 (8%) | 63/705 (9%) | 0.63 |
| ***Prevalence of STI/RTIs*** | | | |
| *Chlamydia trachomatis*, n/N (%) |  |  | 0.29 |
| Negative | 655/703 (93%) | 1412/1497 (94%) |  |
| Positive | 48/703 (7%) | 85/1497 (6%) |  |
| *Neisseria gonorrhea*, n/N (%) |  |  | 0.61 |
| Negative | 683/703 (97%) | 1460/1497 (98%) |  |
| Positive | 20/703 (3%) | 37/1497 (2%) |  |
| *Trichomonas vaginalis*, n/N (%) |  |  | <0.001 |
| Negative | 625/703 (89%) | 1412/1497 (94%) |  |
| Positive | 78/703 (11%) | 85/1497 (6%) |  |
| Bacterial vaginosis, n/N (%) |  |  | <0.001 |
| Negative | 449/695 (65%) | 718/1526 (47%) |  |
| Positive | 246/695 (35%) | 808/1526 (53%) |  |
| Group B *Streptococcus*, n/N (%) |  |  | 0.50 |
| Negative | 559/615 (91%) | 633/705 (90%) |  |
| Positive | 56/615 (9%) | 72/705 (10%) |  |
| ***Delivery details*** | | | |
| Location of delivery, n/N (%) |  |  | <0.001 |
| Study clinic | 694/705 (98%) | 1657/1833 (90%) |  |
| At home or another clinic | 11/705 (2%) | 176/1833 (10%) |  |
| Gestational age at delivery in weeks, median (IQR) | 40 (39-40) | 39 (39-40) | <0.001 |
| Miscarriage, n/N (%) |  |  | 0.003 |
| Yes | 702/705 (100%) | 1795/1833 (98%) |  |
| No | 3/705 (0%) | 38/1833 (2%) |  |
| Preterm delivery*, n/N (%) |  |  | <0.001 |
| ≤37 gestational weeks | 679/705 (96%) | 1679/1833 (92%) |  |
| >37 gestational weeks | 26/705 (4%) | 154/1833 (8%) |  |
| Term low birthweight^, n/N (%) |  |  | 0.68 |
| ≥2500 grams and ≥37 gestational weeks | 649/678 (96%) | 1592/1670 (95%) |  |
| <2500 grams and ≥37 gestational weeks | 29/678 (4%) | 78/1670 (5%) |  |
| Low birthweight^, n/N (%) |  |  | 0.027 |
| <2500 grams | 653/690 (95%) | 1627/1767 (92%) |  |
| ≥2500 grams | 37/690 (5%) | 140/1767 (8%) |  |

Abbreviations: BMI = body mass index; DP = dihydroartemisinin-piperaquine; IQR = interquartile range; MUAC = mid-upper arm circumference; SP = sulfadoxine-pyrimethamine

^#^ Defined as any prior preterm delivery, spontaneous abortion, or stillbirth.

* Includes fetal loss

^ Birthweight outcomes assessed only among live births; term low birthweight excludes preterm births.

**Supplementary Appendix 2.** Description of sensitivity analyses to account for potential bias due to differential timing of swab collection.

**Rationale.** Our primary aim was to determine whether monthly IPTp with SP reduces the risk of STIs/RTIs. While efforts were made to collect vaginal swab samples prior to delivery, 59% (n=1509) of samples collected were collected postpartum, and 2% (n=51) at an unknown date. Of those collected postpartum, the mean time from delivery to collection was 28 days (standard deviation: 5) (see Figure below). Results from samples collected postpartum may not accurately capture infections that occurred during pregnancy, as they may reflect postpartum incident infections or changes in bacterial colonization. This is particularly relevant for bacterial vaginosis, as the vaginal microbiome undergoes substantial shifts in the postpartum period, transitioning to a more *Lactobacillus*-depleted state—a common indicator of bacterial vaginosis.^1,2^

**Figure 1.** Histogram of days since delivery to swab collection among those whose samples were collected postpartum.

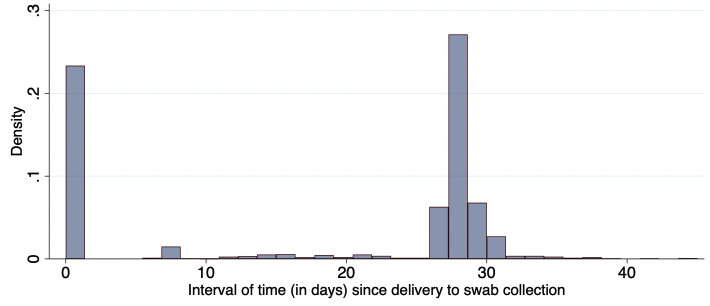

Therefore, we were concerned that failing to account for the timing of swab collection could lead to differential misclassification of STI/RTI status at delivery, whereas restricting analyses to delivery samples alone could potentially introduce selection bias. Given we did not know a plausible range of specificity and sensitivity values, we did not conduct probabilistic bias analyses. However, restricting analyses would not only cause a loss to statistical power, but there was concern that the subgroup with available postpartum samples may not be exchangeable with participants whose samples were collected during pregnancy. If the probability of having a postpartum sample was caused (either directly or indirectly) by the exposure (IPTp-SP) and outcome (STI/RTI status), standard analyses could yield a biased estimate of the treatment effect. Indeed, comparison of participant characteristics by timing of sample collection revealed systematic differences between the two subgroups, including differences in IPTp assignment, STI/RTI prevalence, and preterm birth risk (see **Supplementary Appendix 1**).

Discussions with the study team indicated that timing of sample collection was largely influenced by the location of delivery, which was strongly associated with preterm birth risk. Women who delivered preterm were more likely to have an unexpected or emergency delivery, limiting the opportunity to collect samples before active labor. As both IPTp and STI/RTI risk have been previously thought to be on the causal pathway of preterm birth,^3,4^ this mechanism of selection could induce collider stratification bias if restricted our analytic sample to those whose samples were collected delivery (i.e., conditioning on sample availability—a descendent of a collider, preterm birth). See DAG below.

**Figure 2.** Directed acyclic graph depicting selection bias due to differential timing of sample collection

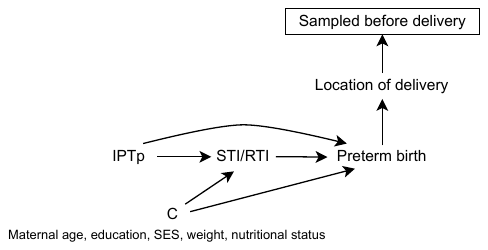

**Abbreviations:** IPTp = intermittent preventive treatment; SES = socioeconomic status; STI/RTI = sexually transmitted infections/reproductive tract infections

To assess the impact of this potential selection bias, we conducted the following sensitivity analyses to test the robustness of our effect estimates. In both approaches, we considered postpartum STI/RTI results as missing and assumed that the missingness was at random, meaning that the observed covariates could account for the underlying missing data mechanism.

**Multiple imputation.** Missing STI/RTI values for individuals without pre-delivery samples (n=1833), along with maternal weight (n=1) and household wealth index (n=14), were imputed using multiple imputation. We used an ensemble machine learning approach (SuperLearner) to flexibly model the relationships between predictors and outcome, allowing for nonlinear relationships and complex interactions between predictors and without requiring strong parametric assumptions. Models included randomized IPTp arm, maternal mid-upper arm circumference, age, weight, household wealth index, current/recent alcohol use, gravidity, preterm delivery status, and prior history of adverse delivery outcome as predictors. For continuous variables, candidate learners included generalized linear models (GLM), predictive mean, and random forest models. For binary variables, candidate learners included logistic regression, GLM with elastic net regularization, and eXtreme gradient boosting. For categorical variables (e.g., wealth status), candidate learners included multinomial logistic regression and random forest algorithms. Ten-fold cross-validation was used to determine the optimal weighted combination of learners. Imputation was performed using the MISL R package^5^ to generate 100 imputed datasets and 20 iterations per dataset. Analytic models were run on each imputed dataset and pooled using Rubin’s combining rules.^6^

**Inverse probability of selection weights (IPSW).** IPSW was used to reweight the participants with non-missing outcomes (i.e., participants with STI/RTI results collected before delivery) to approximate the distribution of the full trial population, as if all participants had samples collected before delivery.^7-9^ Logistic regression was used to estimate the probability of having a sample collected before delivery, using IPTp arm, maternal mid-upper arm circumference, age, and weight at enrollment, household wealth index, gravidity, current/recent alcohol use, history of adverse birth outcome, and preterm birth status as covariates. MUAC, age, weight, and gravidity were modeled as restricted cubic splines with five knots. The inverse of these estimated probabilities was then used as weights in the final analytic models. Weights were stabilized by multiplying them by the marginal probability of having a delivery sampling (705/2538=28%) (see equations below):

$$\boldsymbol{Selection model:}$$

$\mathrm{logit} \left( \hat{P}\left( S_{i}=1 \right) \right)= \beta_{0}+\beta_{1}{IPTp}_{i}+\beta_{2}Wealth+\beta_{3}BMI+\beta_{4}Education+\beta_{5}MUAC+\beta_{6}Age+\beta_{7}Weight+\beta_{8}Gravidity+ \beta_{9}Alcohol use+{\beta_{10}History of adverse birth outcome+\beta}_{11}Preterm$

where $S_{i}=1$ if participant $i$ had a sample collected before delivery and $and S_{i}=0$ otherwise. $MUAC$, $Age$, and $Weight$ were modeled as restricted cubic splines with five knots.

$$\boldsymbol{Stabilized weights:}$$

$$w_{i}^{stablized}=\left\{ \begin{aligned} \frac{\hat{P}\left( S_{i}=1 \right)}{\hat{P}\left( S_{i}=1 | IPTp,STI, Preterm, C, E \right)} &if S_{i}=1 \\ 0 &if S_{i}=0 \end{aligned} \right.$$

where $S_{i}$ is an indicator of selection into the analytic sample, $C$ denotes confounders of the relationship between STI and adverse birth outcomes, and $E$includes prognostic variables (i.e., gravidity and prior history of adverse birth outcomes).

Like all causal inference methods, IPSW relies on the assumptions of exchangeability, consistency, and positivity. To assess potential violations of the positivity assumption (i.e., whether all individuals had a non-zero probability of having a sample collected before delivery given their covariate distribution), we examined the distribution of estimated probabilities using a kernel density plot. Based on the plot below, a subset of individuals showed poor overlap in selection probabilities (stabilized weights >2), indicating potential violations of the positivity assumption. To address this, weights were truncated at 2 to reduce instability and improve the reliability of effect estimates. As a result, the IPSW estimates should be interpreted as applying to the subset of the population with adequate covariate overlap, rather than the full trial population.

**Figure 3.** Kernel density plot to assess for potential positivity violations in IPSW estimation.

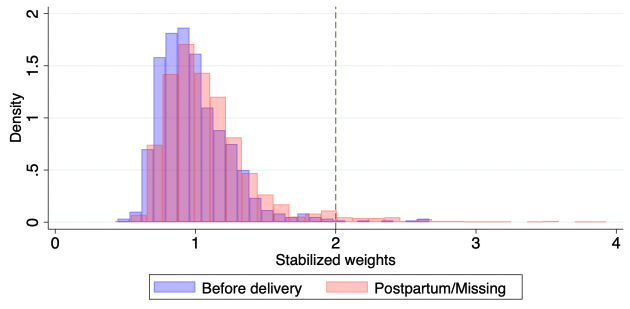

**Supplementary Appendix 3.** Results from sensitivity analyses comparing STI/RTI risk between IPTp arms

**Figure 1**. Comparison of IPTp-SP alone vs. IPTp-DP alone

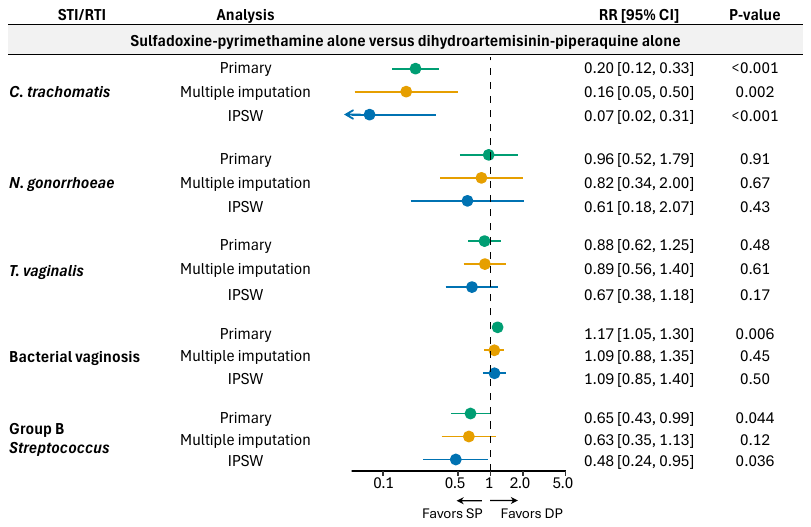

Abbreviations: *C. trachomatis* = *Chlamydia trachomatis*; DP = dihydroartemisinin-piperaquine; IPSW = inverse probability of selection weights; *N. gonorrhoeae* = *Neisseria gonorrhoeae*; RR = relative risk ratio; SP = sulfadoxine-pyrimethamine; *T. vaginalis* = *Trichomonas vaginalis*

**Figure 2**. Comparison of IPTp-DP+SP vs. IPTp-DP alone

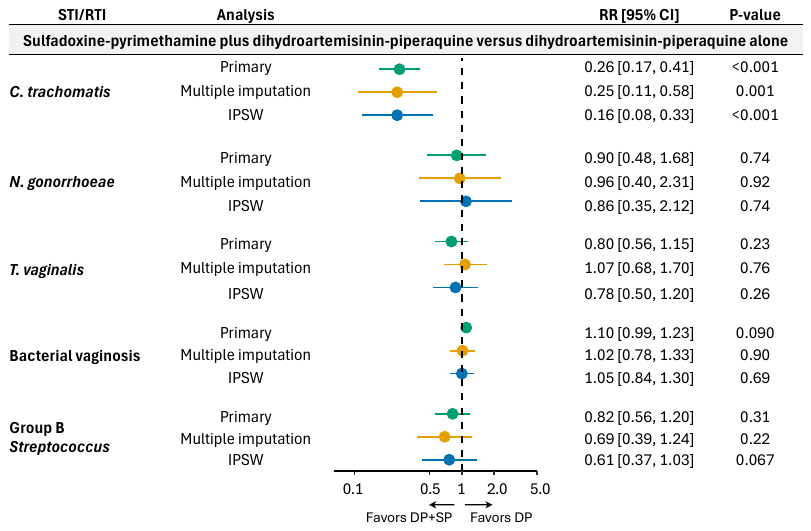

Abbreviations: *C. trachomatis* = *Chlamydia trachomatis*; DP = dihydroartemisinin-piperaquine; IPSW = inverse probability of selection weights; *N. gonorrhoeae* = *Neisseria gonorrhoeae*; RR = relative risk ratio; SP = sulfadoxine-pyrimethamine; *T. vaginalis* = *Trichomonas vaginalis*

**Figure 3**. Comparison of any SP-containing IPTp regimen vs. IPTp-DP alone

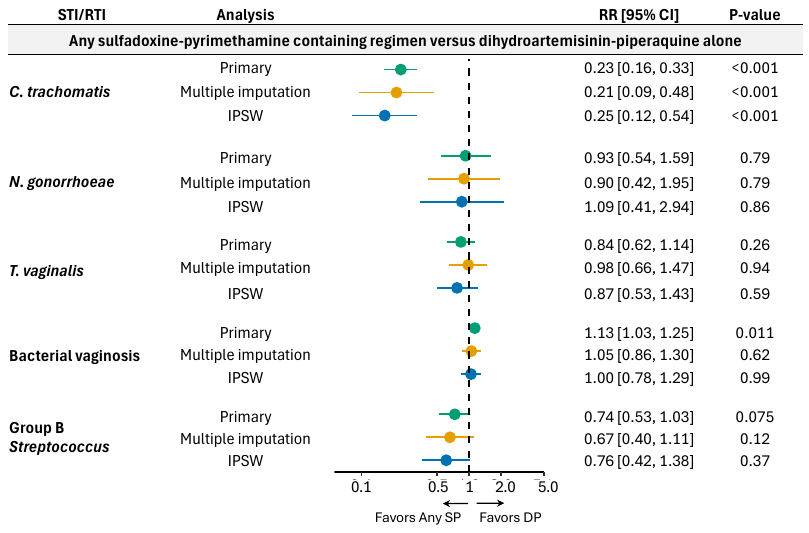

Abbreviations: *C. trachomatis* = *Chlamydia trachomatis*; DP = dihydroartemisinin-piperaquine; IPSW = inverse probability of selection weights; *N. gonorrhoeae* = *Neisseria gonorrhoeae*; RR = relative risk ratio; SP = sulfadoxine-pyrimethamine; *T. vaginalis* = *Trichomonas vaginalis*

**Supplementary Appendix 5.** Results from sensitivity analyses estimating effect of STI/RTI risk on adverse birth outcomes

**Figure 1.** Effects of STIs/RTIs on preterm delivery

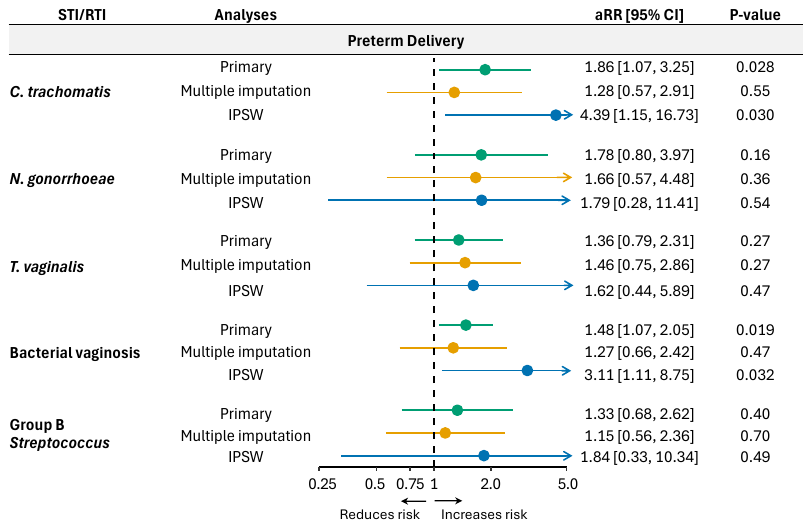

Abbreviations: aRR = adjusted relative risk ratio; *C. trachomatis* = *Chlamydia trachomatis*; IPSW = inverse probability of selection weights; *N. gonorrhoeae* = *Neisseria gonorrhoeae*; STI/RTI = sexually transmitted infections/reproductive tract infections; *T. vaginalis* = *Trichomonas vaginalis*

**Figure 2.** Effects of STIs/RTIs on term low birthweight

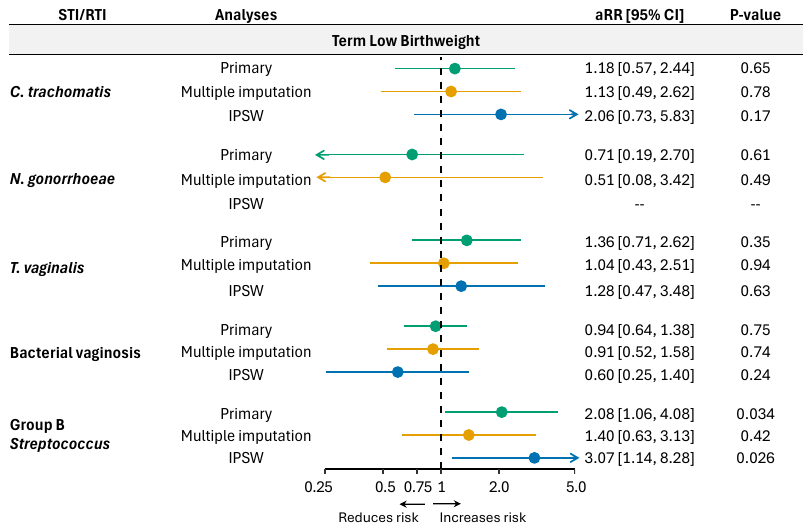

Abbreviations: aRR = adjusted relative risk ratio; *C. trachomatis* = *Chlamydia trachomatis*; IPSW = inverse probability of selection weights; *N. gonorrhoeae* = *Neisseria gonorrhoeae*; STI/RTI = sexually transmitted infections/reproductive tract infections; *T. vaginalis* = *Trichomonas vaginalis*

**Figure 3.** Effects of STIs/RTIs on low birthweight

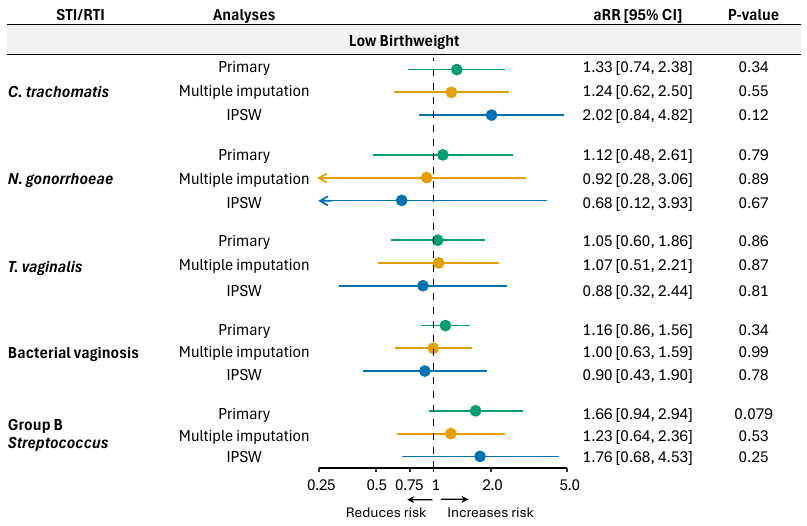

Abbreviations: aRR = adjusted relative risk ratio; *C. trachomatis* = *Chlamydia trachomatis*; IPSW = inverse probability of selection weights; *N. gonorrhoeae* = *Neisseria gonorrhoeae*; STI/RTI = sexually transmitted infections/reproductive tract infections; *T. vaginalis* = *Trichomonas vaginalis*

**Figure 4.** Effects of STIs/RTIs on small-for-gestational age

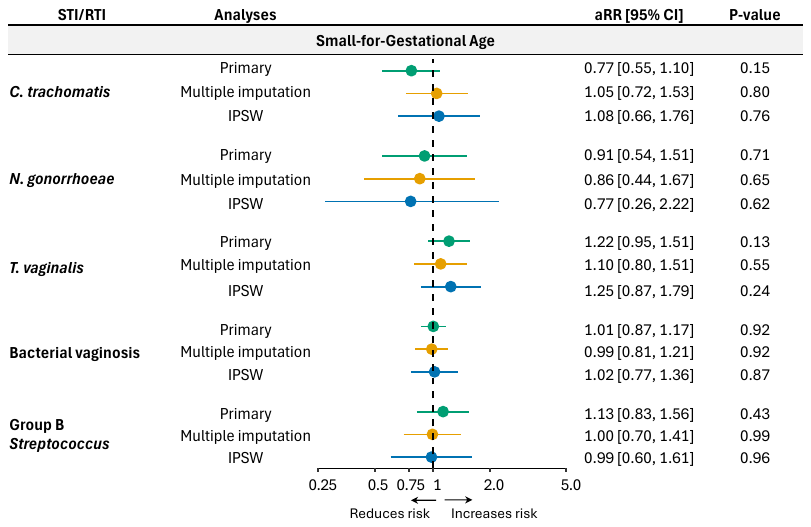

Abbreviations: aRR = adjusted relative risk ratio; *C. trachomatis* = *Chlamydia trachomatis*; IPSW = inverse probability of selection weights; *N. gonorrhoeae* = *Neisseria gonorrhoeae*; STI/RTI = sexually transmitted infections/reproductive tract infections; *T. vaginalis* = *Trichomonas vaginalis*
